# Assessing ChatGPT’s Mastery of Bloom’s Taxonomy using psychosomatic medicine exam questions

**DOI:** 10.1101/2023.08.18.23294159

**Authors:** Anne Herrmann-Werner, Teresa Festl-Wietek, Friederike Holderried, Lea Herschbach, Jan Griewatz, Ken Masters, Stephan Zipfel, Moritz Mahling

## Abstract

**Introduction:** Large language models (LLMs) such as GPT-4 are increasingly used in medicine and medical education. However, these models are prone to “hallucinations” – outputs that sound convincing while being factually incorrect. It is currently unknown how these errors by LLMs relate to the different cognitive levels defined in Bloom’s Taxonomy.

**Methods:** We used a large dataset of psychosomatic medicine multiple-choice questions (MCQ) (N = 307) with real-world results derived from medical school exams. GPT-4 answered the MCQs using two distinct prompt versions – detailed and short. The answers were analysed using a quantitative and qualitative approach. We focussed on incorrectly answered questions, categorizing reasoning errors according to Bloom’s Taxonomy.

**Results:** GPT-4’s performance in answering exam questions yielded a high success rate: 93% (284/307) for the detailed prompt and 91% (278/307) for the short prompt. Questions answered correctly by GPT-4 had a statistically significant higher difficulty compared to questions that GPT-4 answered incorrectly (p=0.002 for the detailed prompt and p<0.001 for the short prompt). Independent of the prompt, GPT-4’s lowest exam performance was 78.9%, always surpassing the pass threshold. Our qualitative analysis of incorrect answers, based on Bloom’s Taxonomy, showed errors mainly in the “remember” (29/68) and “understand” (23/68) cognitive levels. Specific issues arose in recalling details, understanding conceptual relationships, and adhering to standardized guidelines.

**Discussion:** GPT-4 displayed a remarkable success rate when confronted with psychosomatic medicine multiple-choice exam questions, aligning with previous findings. When evaluated against Bloom’s hierarchical framework, our data revealed that GPT-4 occasionally ignored specific facts (“remember”), provided illogical reasoning (“understand”), or failed to apply concepts to a new situation (“apply”). These errors, though confidently presented, could be attributed to inherent model biases and the tendency to generate outputs that maximize likelihood.

**Conclusion:** While GPT-4 mostly excels at medical exam questions, discerning its occasional cognitive errors is crucial.

## 1 Introduction

The recent developments in artificial intelligence (AI) hold transformative potential for various fields, including medicine^1^ and medical education.^2^ In November 2022, OpenAI launched GPT-3, a large language model (LLM).^3^ Its high-quality performance surprised even experts and led to huge public interest. This is particularly true for school and higher education settings, where GPT-3 prompted manifold discussions on potential benefits and harms.^4,5^ In medical education, LLMs have the potential to revolutionize the current teaching approaches and thus ultimately improve physician performance and healthcare outcomes. However, before LLMs are deeply integrated into medical education, their performance must be comprehensively assessed in the context of medical education. It is especially critical to evaluate the capabilities of AI and LLMs within the educational theoretical frameworks.

One of the most-used frameworks in medical education is Benjamin Bloom’s taxonomy^6,7^ of learning outcomes. Briefly, Bloom and subsequent colleagues developed a hierarchical classification of cognitive processes, ordered from lower-order cognitive skills, such as knowledge recall (“remember”) and comprehension (“understand”), to higher-order thinking such as application (“apply”), analysis (“analyse”), evaluation (“evaluate”) and creation (“create”).^8^ From its first publication in 1956 until now, this taxonomy has been used as a common language for educational instructors and still influences the field of medical education.^8^

Although derived from human learning processes, the taxonomy developed by Bloom provides an ideal framework to describe cognitive processes that underlay success and failure. Recently, LLMs have been assessed for their (mostly surprisingly good) performance in various fields of medicine, ranging from specific subjects to board exams.^9–11^ However, the errors that LLMs make have not been evaluated in detail. For example, while LLMs might successfully recall facts (“remember”), they might struggle to apply those facts to a different context (or vice versa). At this stage, the authors acknowledge that applying terms like “remember” and “struggle” are anthropomorphisms for ease of reading, as an LLM currently does neither and merely generates responses based on language usage statistical probabilities, using a “next-word prediction paradigm”.^12^

Our intention was thus to use Bloom’s Taxonomy in order to gain a better understanding of the failures of LLMs. For studies of human medicine and the aforementioned use cases for LLMs, multiple-choice questions (MCQ) remain a primary written exam form and are used for summative and formative assessments.^13^ In Bloom’s Taxonomy, MCQs are often used to assess lower-order cognitive skills, such as knowledge recall (“remember”) and comprehension (“understand”), but may also probe higher-order thinking such as application (“apply”), analysis (“analyse”) and evaluation (“evaluate”).^14^ The form of MCQs thus offers a good window for evaluating different cognitive processes.

To elucidate cognitive processes and correct/incorrect reasoning, a medical field that heavily relies on language and factual understanding is instrumental. Given the interplay of psychological, social, and biological factors, psychosomatic medicine presents such a challenge. The field’s heavy reliance on verbal and written communication for diagnosis and treatment makes it particularly challenging. Additionally, the combination of strict diagnostic criteria with a nuanced understanding of the patient’s language positions it as an ideal testing ground for the capabilities of language models.

This paper presents a mixed-methods study designed to explore how GPT-4 performs (and fails) with regard to Bloom’s Taxonomy. First, we assessed the performance of GPT-4 in a large set of psychosomatic medicine exam questions and compared the results to responses from our cohort of medical students, providing a human comparison and quality indicators. For a deeper understanding of the results, we employed qualitative methods to understand the model’s performance and to assess the strengths and weaknesses of LLMs in relation to Bloom’s Taxonomy. The findings of this study will provide critical insights into the practical applications and limitations of LLMs, such as GPT-4, in medical education.

## 2 Methods

### 2.1 Exams

A total of 16 examinations from winter term 2014/2015 to summer term 2022 were retrieved from the Department of Psychosomatic Medicine and Psychotherapy faculty’s online exam programme integrated management system (IMS) (https://www.ucan-assess.org/ims/). In addition to question stems, answers, and distractors, the system also offers quality criteria for each individual question (discriminatory power and level of difficulty).

Each examination consists of 20 MCQs, each with one answer and four distractors. Diagnostic and therapeutic questions cover topics concerning anxiety disorders, depression, eating, and somatoform and trauma disorders. Questions concerning motivational interviewing techniques are also part of the examination.

Undergraduate medical students take this examination at the end of their third year after having attended 7.5 hours of lectures and 18 hours of practical classes on psychosomatic medicine and psychotherapy.

To pass, a student is usually required to correctly answer 12 out of 20 questions; an adjustment of this passing score is possible if, for example, a question has a too-poor performance.

We assessed the questions for their suitability to be analysed by GPT-4. From a total of 320 questions, 13 (4%) were excluded. Excluded were questions that were not single choice (n = 6), ambiguous questions (n = 3), questions featuring a graphic that had to be analysed (n = 2), and questions that covered a case represented in multiple single questions (n = 2).

We used GPT-4 to answer every single question (model “gpt-4”, OpenAI LP, San Francisco, California, USA). For each question, we generated a detailed and a short prompt version. The prompts were created by the authors in an iterative process using the web-interface ChatGPT Plus to achieve ideal performance. The most relevant difference between versions was that the detailed version included the command to critically reflect upon the answers and justify the choice made. For a detailed example, please see Table 1. We used the application programming interface (API) provided by OpenAI to post the question to GPT-4 and retrieve the answer. While all data are presented in English, the data acquisition was performed in German.

**Table 1:**
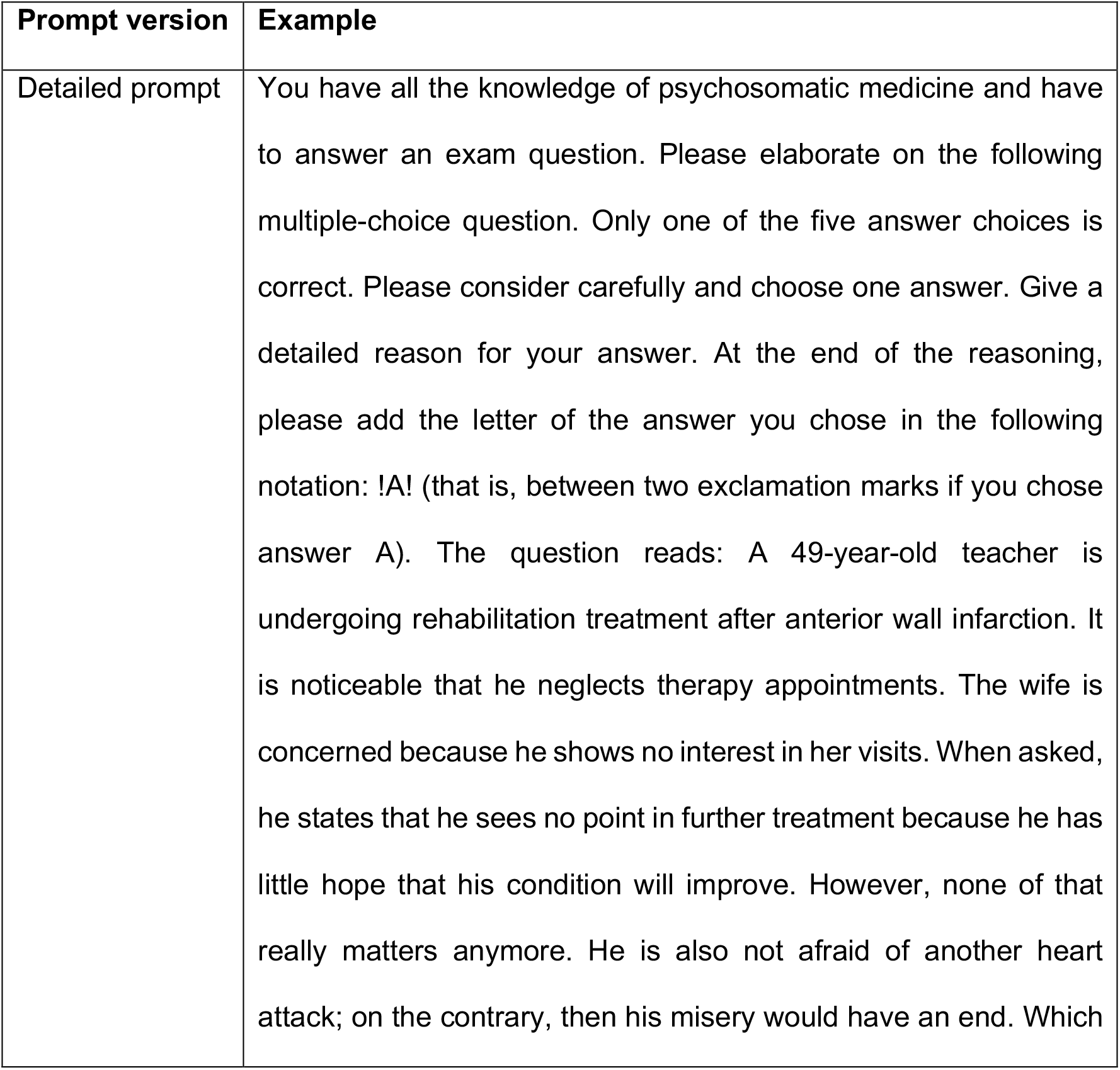
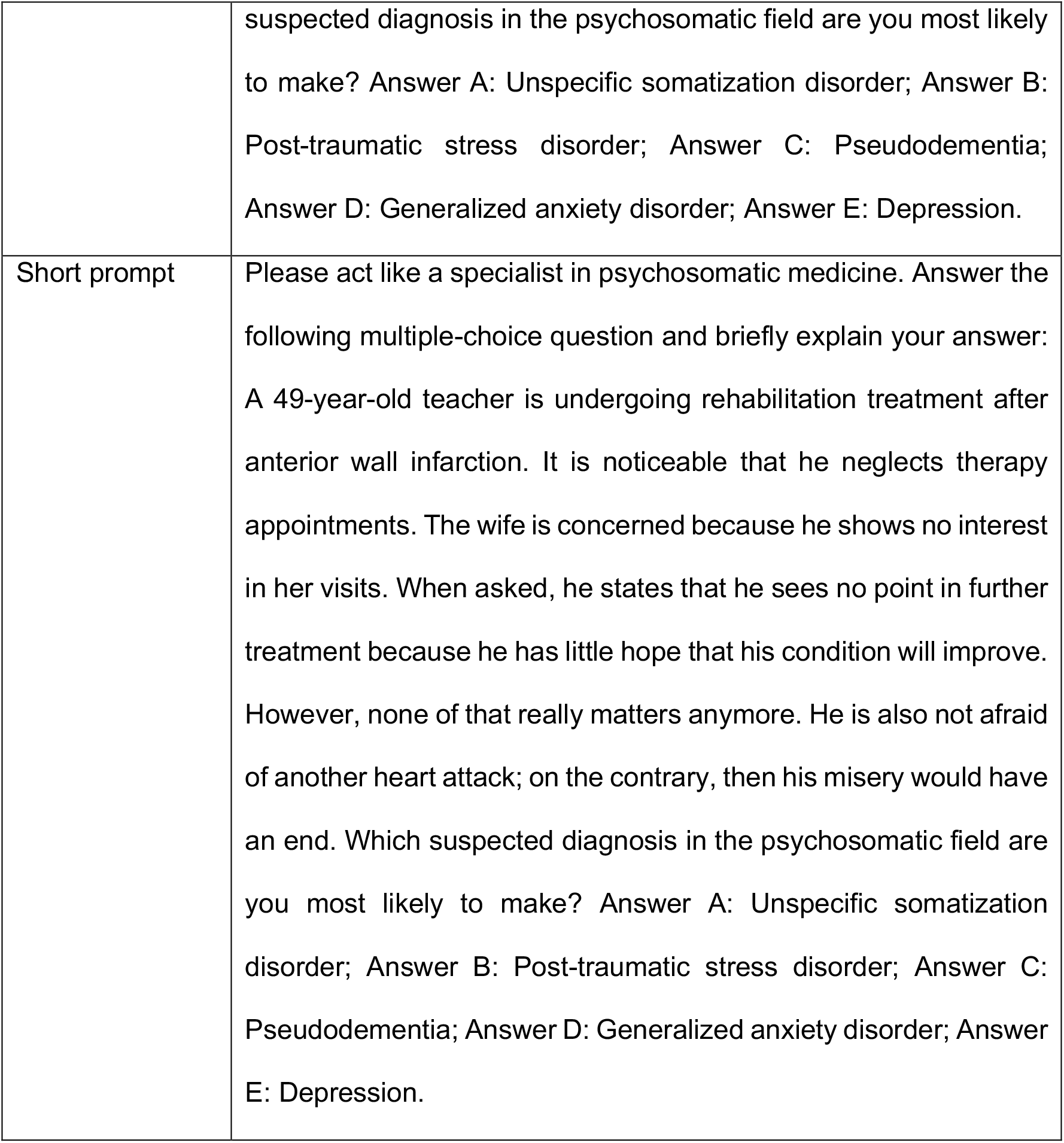
Examples of detailed and short prompt versions.

| Prompt version | Example |
| --- | --- |
| Detailed prompt | You have all the knowledge of psychosomatic medicine and have to answer an exam question. Please elaborate on the following multiple-choice question. Only one of the five answer choices is correct. Please consider carefully and choose one answer. Give a detailed reason for your answer. At the end of the reasoning, please add the letter of the answer you chose in the following notation: !A! (that is, between two exclamation marks if you chose answer A). The question reads: A 49-year-old teacher is undergoing rehabilitation treatment after anterior wall infarction. It is noticeable that he neglects therapy appointments. The wife is concerned because he shows no interest in her visits. When asked, he states that he sees no point in further treatment because he has little hope that his condition will improve. However, none of that really matters anymore. He is also not afraid of another heart attack; on the contrary, then his misery would have an end. Which |
|  | suspected diagnosis in the psychosomatic field are you most likely to make? Answer A: Unspecific somatization disorder; Answer B: Post-traumatic stress disorder; Answer C: Pseudodementia; Answer D: Generalized anxiety disorder; Answer E: Depression. |
| Short prompt | Please act like a specialist in psychosomatic medicine. Answer the following multiple-choice question and briefly explain your answer:<br>A 49-year-old teacher is undergoing rehabilitation treatment after anterior wall infarction. It is noticeable that he neglects therapy appointments. The wife is concerned because he shows no interest in her visits. When asked, he states that he sees no point in further treatment because he has little hope that his condition will improve. However, none of that really matters anymore. He is also not afraid of another heart attack; on the contrary, then his misery would have an end. Which suspected diagnosis in the psychosomatic field are you most likely to make? Answer A: Unspecific somatization disorder; Answer B: Post-traumatic stress disorder; Answer C: Pseudodementia; Answer D: Generalized anxiety disorder; Answer E: Depression. |

### 2.2 Data analysis

The responses given by GPT-4 were compared to the answer indicated by the answer index (e.g., “A” or “C”) and stored in Microsoft Excel (Version 16.0.10394.20022).

#### 2.2.1 Quantitative data analysis

Quantitative analyses and figure generation were performed using R Statistical software (R version 4.3.1).^15^ Briefly, we combined all tables with relevant data, i.e., from GPT-4 and the aggregated results of the students’ exams (such as item difficulty). For each prompt version, we analysed the ratio of correctly and incorrectly answered questions. We further compared the question difficulty (from the aggregated student data) between questions answered correctly and incorrectly by GPT-4. The difficulty of a question can be understood as the fraction of students answering a question correctly, with 0 representing a very difficult question and 1 a very easy question.^16^ To test for statistical significance, a Wilcoxon rank sum test was used. A p-level < 0.05 was considered statistically significant. If not stated otherwise, medians and 25%/75% quartiles are given (Q25–Q75) in the results.

#### 2.2.2 Qualitative data analysis

Two authors (TFW, FH) separately coded each text response. The answers from GPT-4 were analysed inductively and iteratively according to Mayring’s qualitative content analysis^17^ and as described previously by our group.^18^ The goal of the analysis were defined according to the answers to the examination questions. As the main category, we used the correct or incorrect answering of the question. We further focussed mainly on incorrect answers.

In the answer text, individual reasoning was categorized according to Bloom’s Taxonomy as revised by Krathwohl.^8^ In a second step, each of the raters coded the answers using MAXQDA (VERBI software, version 12.3.2). In order to obtain the same level of abstraction in building the categories, the raters revised them together and agreed on the final categories by paraphrasing representative examples and building a hierarchy of categories. Following this, both raters independently worked through the material again. When GPT-4’s responses were wrong, the explanation was analysed using Bloom’ s Taxonomy levels, including remember, understand, apply, analyse, evaluate, and create.^8^

### 2.3 Ethics

The Ethics Committee of the Faculty of Medicine at University Hospital Tübingen approved the study (number 076/2023A). All data were kept anonymous and were not associated with students.

## 3 Results

### 3.1 Quantitative Results

#### 3.1.1 Distribution of correctly and incorrectly answered questions

For the detailed prompt, the success rate was 93% (284 of 307 questions). For the short prompt, GPT-4 answered 91% of the questions correctly (278 of 307 questions). The distribution is shown in Figure 1.

**Figure 1:**
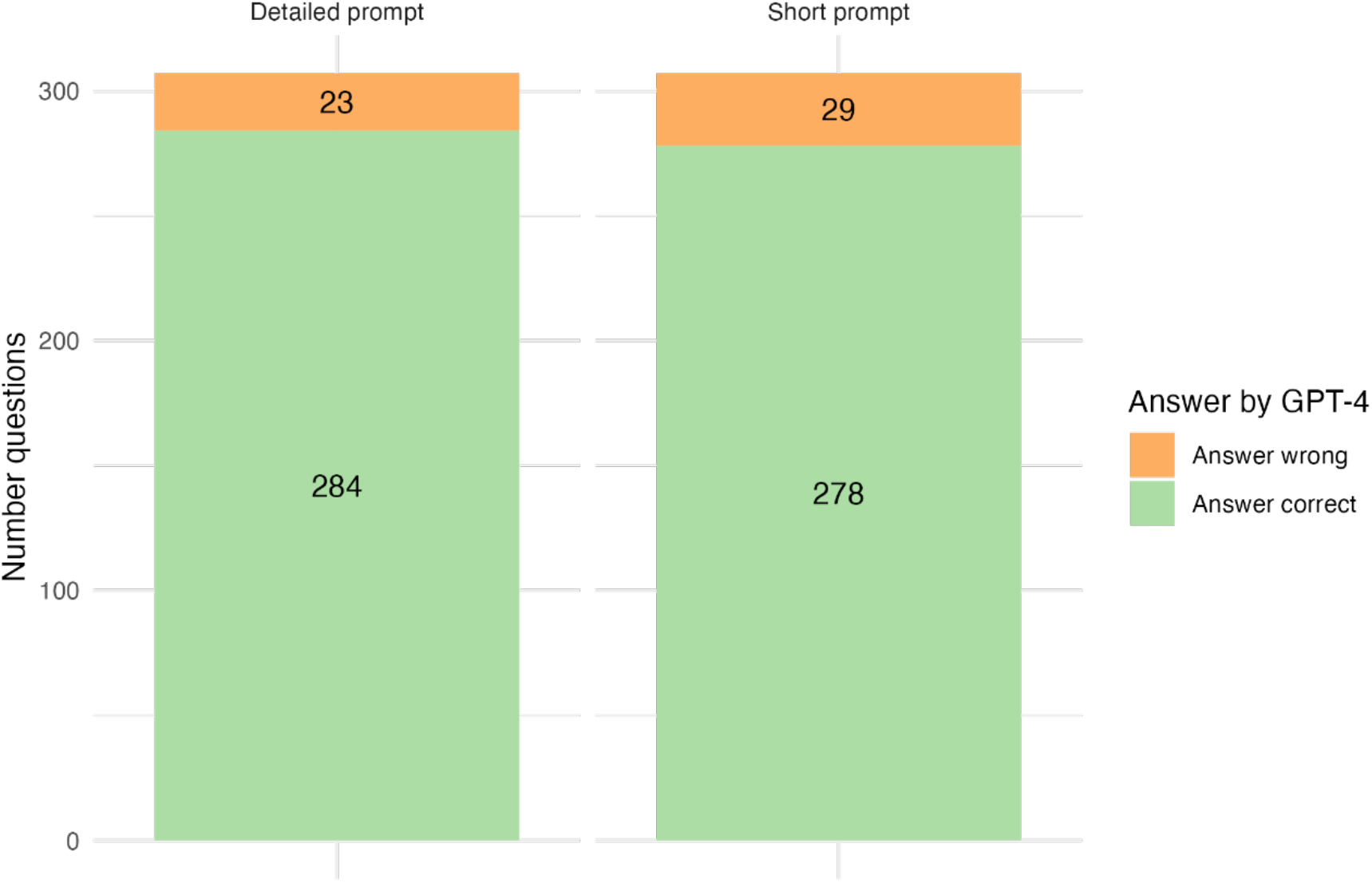
Distribution of correctly and incorrectly answered questions by prompt version.

#### 3.1.2 Question difficulty

Among all questions, the median difficulty was 0.892 (Q25-Q75: 0.705–0.949). The distribution of the question difficulty for correctly and incorrectly answered questions is displayed in Figure 2.

**Figure 2:**
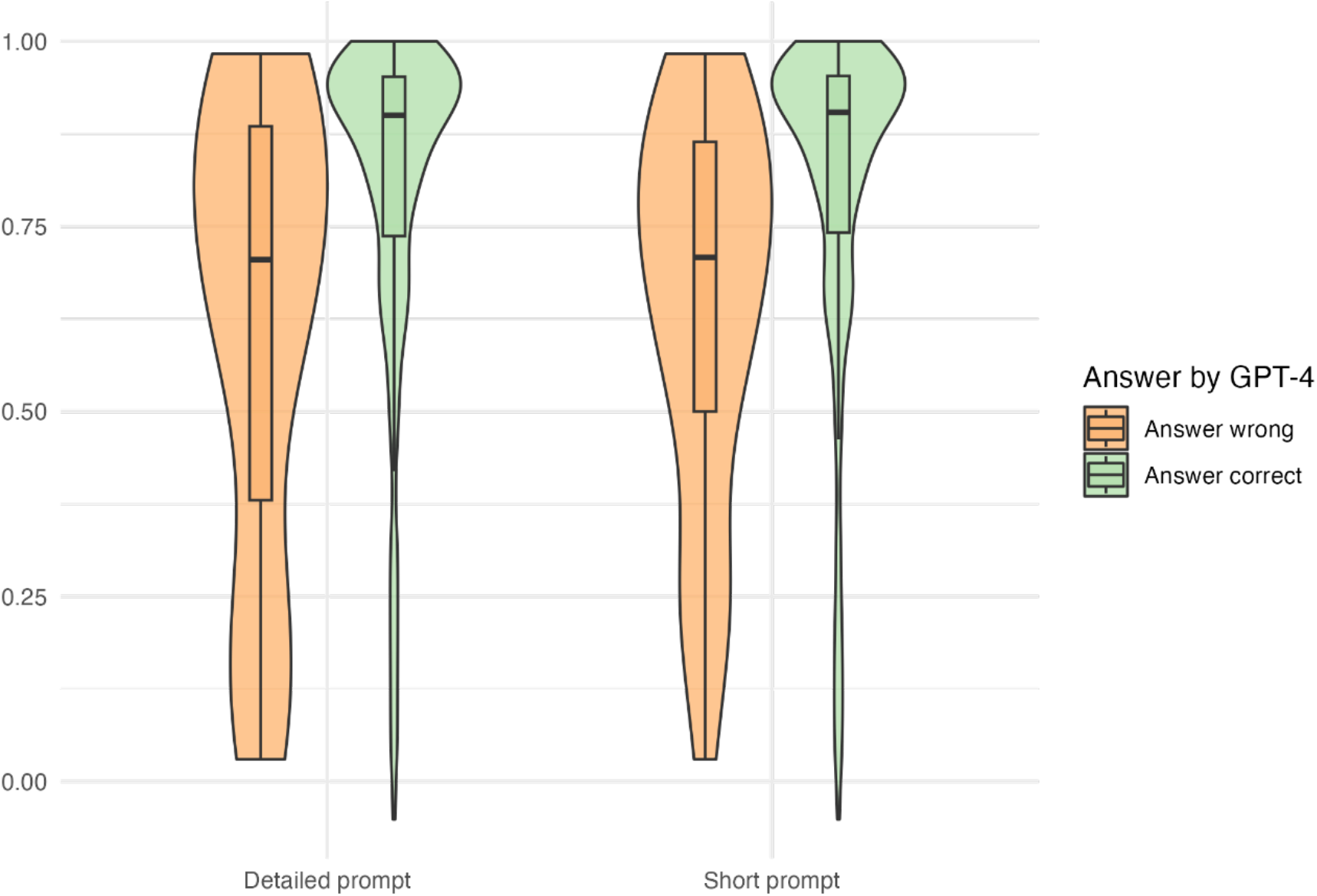
Question difficulty by prompt version and answer correctness.

**Figure 3:**
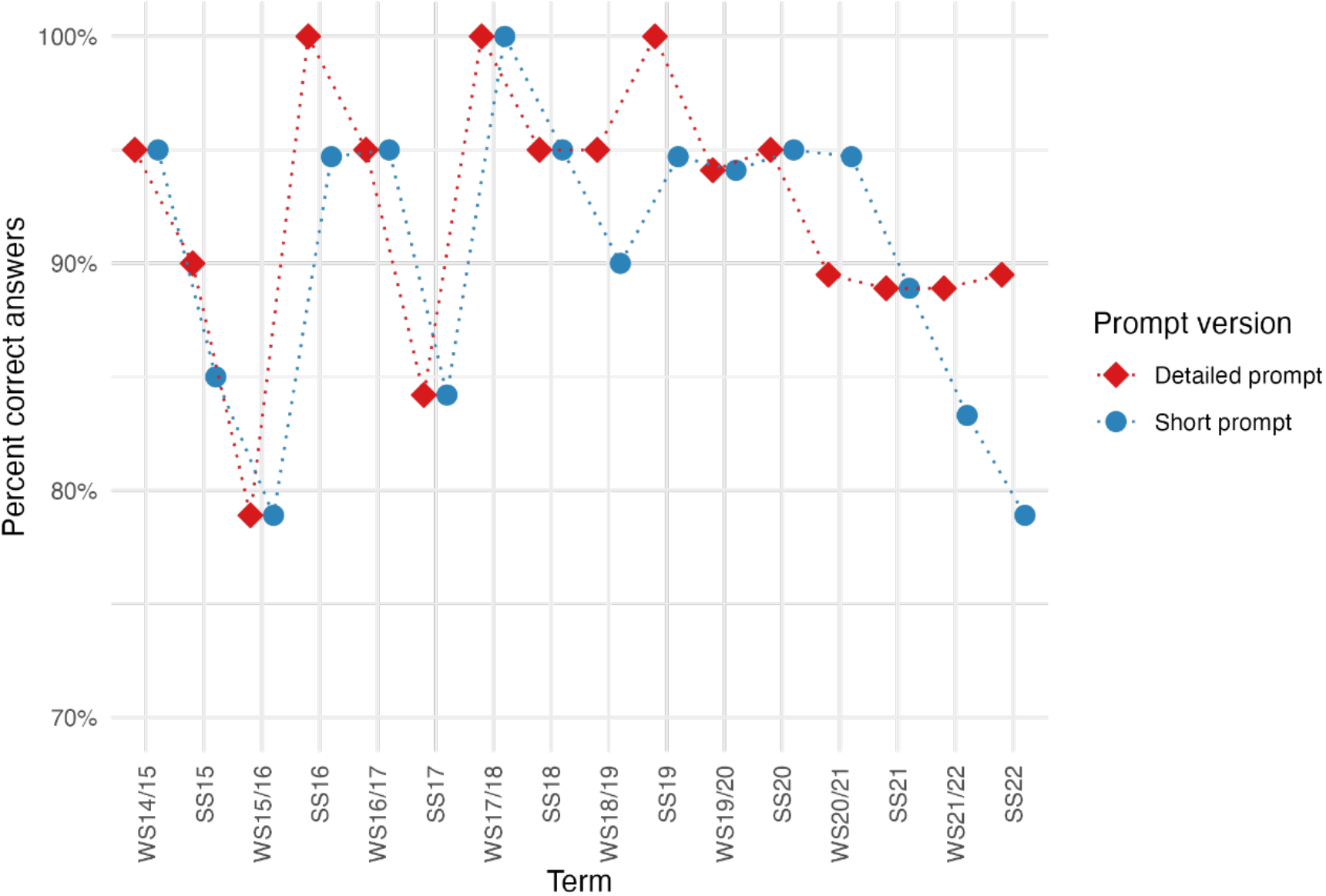
Performance of GPT-4 for all relevant terms for the detailed prompt (red) and short prompt (blue).

For the detailed prompt, questions answered correctly had a higher difficulty (0.900, Q25-Q75: 0.737–0.952) when compared to questions answered incorrectly (0.705, Q25-Q75: 0.380–0.885). This difference was statistically significant (p = 0.002).

When analysing the short prompt version, we also found a lower difficulty among incorrectly answered questions (0.708, Q25-Q75: 0.500–0.864) when compared to correctly answered questions (0.904, Q25-Q75: 0.741–0.953). A significant difference was detected between the correctly and incorrectly answered questions (p < 0.001).

#### 3.1.3 Exam scores

We further analysed the performance of GPT-4 for all 16 individual terms/exams (Table 1). Regardless of the prompt version, GPT-4 never performed below 78.9% and thus always passed the exams. A total of three exams (one exam for both prompt versions and two exams in the detailed prompt version only) were passed with a score of 100%. The exam rates are displayed in Table 2.

**Table 2:** Results of GPT-4 for all relevant terms (WS: winter term, SS: summer term).

| Term | Questions:<br>Total<br>included | Short prompt |  |  | Detailed prompt |  |  |
| --- | --- | --- | --- | --- | --- | --- | --- |
|  |  | Answer<br>correct | Answer<br>incorrect | Percent<br>correct | Answer<br>correct | Answer<br>incorrect | Percent<br>correct |
| **WS14/15** | 20 | 19 | 1 | 0.950 | 19 | 1 | 0.950 |
| **SS15** | 20 | 17 | 3 | 0.850 | 18 | 2 | 0.900 |
| **WS15/16** | 19 | 15 | 4 | 0.789 | 15 | 4 | 0.789 |
| **SS16** | 19 | 18 | 1 | 0.947 | 19 | 0 | 1.000 |
| **WS16/17** | 20 | 19 | 1 | 0.950 | 19 | 1 | 0.950 |
| **SS17** | 19 | 16 | 3 | 0.842 | 16 | 3 | 0.842 |
| **WS17/18** | 20 | 20 | 0 | 1.000 | 20 | 0 | 1.000 |
| **SS18** | 20 | 19 | 1 | 0.950 | 19 | 1 | 0.950 |
| **WS18/19** | 20 | 18 | 2 | 0.900 | 19 | 1 | 0.950 |
| **SS19** | 19 | 18 | 1 | 0.947 | 19 | 0 | 1.000 |
| **WS19/20** | 17 | 16 | 1 | 0.941 | 16 | 1 | 0.941 |
| **SS20** | 20 | 19 | 1 | 0.950 | 19 | 1 | 0.950 |
| **WS20/21** | 19 | 18 | 1 | 0.947 | 17 | 2 | 0.895 |
| **SS21** | 18 | 16 | 2 | 0.889 | 16 | 2 | 0.889 |
| **WS21/22** | 18 | 15 | 3 | 0.833 | 16 | 2 | 0.889 |
| **SS22** | 19 | 15 | 4 | 0.789 | 17 | 2 | 0.895 |

### 3.2 Qualitative results

After the quantitative results were obtained, we analysed the answers given by GPT-4 in detail by applying content analysis.

#### 3.2.1 Correctly answered questions

When GPT-4 answered a question correctly, it provided either a detailed or short explanation (Table 3). In the detailed version, GPT-4 also commented on the distractors. For correctly answered questions, the content analysis revealed logical reasoning and correct utilization of facts. No answers were found to be either implausible or incorrect regarding the facts cited.

**Table 3:** (Correct) Answers by GPT-4 for the question described in Table 1 for the detailed and short prompt version.

|  |  |
| --- | --- |
| Detailed prompt | Explanation: After careful consideration of the various answer options and taking into account all the knowledge of psychosomatic medicine, answer E: Depression is the most plausible.<br>In this case, the 49-year-old teacher shows clear signs of emotional impairment after an anterior wall infarction. These include disinterest in therapy appointments, lack of interest in his wife's visits, hopelessness about his recovery, and no sense of fear of another heart attack – on the contrary, he sees the prospect of an end to his "misery" as a positive thing. All these signs point to a possible depressive episode.<br>Answer A: Nonspecific somatization disorder can be ruled out because the patient does not have a multitude of physical symptoms without a demonstrable organic cause.<br>Answer B: Post-traumatic stress disorder could be considered due to the event of the infarction, but the typical symptoms such as memories or flashbacks of the traumatic event, avoidance behaviour, and increased irritability are missing in the description of the case.<br>Answer C: Pseudodementia refers to cognitive impairment caused by depressive symptoms rather than actual dementia. Although this would be possible in the case of depression, the question does not provide enough information about cognitive impairment to support this diagnosis.<br>Answer D: Generalized Anxiety Disorder does not fit the case as the patient states that he is not afraid of having another heart attack and overall does not show excessive fear or anxiety.<br>Therefore, answer E: Depression is the most plausible tentative diagnosis in the psychosomatic field in this case. |
| Short prompt | Answer E: Depression<br>The symptoms described, such as lack of interest in further treatment, lack of hope for improvement, dejection and even death wishes, most likely indicate depression. The patient shows signs of anhedonia (inability to feel pleasure or interest in activities) and negative thought patterns. In such cases, psychosomatic assessment and appropriate treatment is important to aid the patient's recovery. |

#### 3.2.2 Incorrectly answered questions

When a question was answered incorrectly by GPT-4, the explanation’s reasoning was analysed based on Bloom’s Taxonomy, including remember, understand, apply, analyse, evaluate, and create (Table 4). The most mistakes were found for “remember” (n = 29), followed by “understand” (n = 23) and “apply” (n = 15). Only one erroneous reasoning example was found for “evaluate”. In the following sections, examples for each category are given in detail.

**Table 4:** Count of incorrect reasoning by GPT-4 for each category of Bloom’s Taxonomy. For this analysis, all answers (detailed and short prompt) were analysed together.

| Category of Bloom's Taxonomy | N total |
| --- | --- |
| Remember | 29 |
| Understand | 23 |
| Apply | 15 |
| Analyse | 0 |
| Evaluate | 1 |
| Create | 0 |
| **Total** | **68** |

##### 3.2.2.1 Bloom’s Taxonomy: Remember

In this section, we provide an example of a question for which GPT-4 failed to remember facts. The questions presented the case of a man with recurrent physical complaints without organic cause. The complaints persisted for ten months, making it appropriate for an “undifferentiated somatoform disorder” (F45.1 according to the International Statistical Classification of Diseases and Related Health Problems 10th Revision (ICD-10)). A “somatization disorder” (F45.0) cannot be chosen, as it requires “symptoms of at least two years duration”.^19^

However, GPT-4 incorrectly chose “somatization disorder”. In its reasoning, GPT-4 stated:

*“… I would choose answer C: Somatization disorder (F45.0) as the most appropriate diagnosis for this patient. Somatization disorder (F45.0) is characterized by persistent, multiple, and recurrent physical complaints for which no sufficient organic cause can be found. The symptoms can occur in different organ or functional systems and are usually severe enough to interfere with the patient’s daily life.”*

In its recall, GPT-4 did not mention the required duration of the symptoms, as stated in the ICD-10. From what can be observed in the answer, the concept used by GPT-4 thus missed at least one important fact, and the failure could thus be attributed to GPT-4’s not “remembering”.

##### 3.2.2.2 Bloom’s Taxonomy: Understand

The second level of Bloom’s Taxonomy covers the understanding of facts, defined as “determining the meaning of instructional messages, including oral, written, and graphic communication”^8^. An example of GPT-4’s not understanding some basic concepts can be found in a question concerning anorexia nervosa. The answer covers the fact that anorexia nervosa can be accompanied by transient hypercholesterolemia that does not necessitate treatment.^20,21^ However, GPT-4 picked one of the distractors. The distractor chosen by GPT-4 states that the basic metabolic rate is increased during phases of low body weight and decreases after refeeding^22^ – but the opposite is true. In its explanation, GPT-4 thought about these facts (and remembered them correctly in sentence 3!) but did not seem to understand how they relate and interact:

*“In patients with anorexia nervosa, the basal metabolic rate is initially elevated at the onset of weight gain and decreases dramatically during refeeding. Food intake should be adjusted accordingly. Basal metabolic rate here refers to the amount of energy the body needs at rest to maintain its basic functions. In people with anorexia nervosa, the body is initially very efficient at obtaining energy due to persistent nutrient deficiency. However, as refeeding progresses, the basal metabolic rate drops dramatically as the body re-learns to store energy.”*

##### 3.2.2.3 Bloom’s Taxonomy: Apply

The third level of Bloom’s Taxonomy refers to the application of previously learned content to other areas. An error in this domain became evident when we asked GPT-4 a question about the diagnosis and treatment of a depressive episode. The answer was that symptoms/criteria must be present for at least 14 days, which is clearly required by ICD-10.^19^ However, GPT-4 rates this answer as incorrect by implying that this time frame might be more of a rough guideline, not a strict minimum requirement.

*“In the diagnosis of depressive episodes, the time criterion is an important factor. However, many experts believe that a period of at least two weeks is rather a rough guideline. Some patients can also go through a depressive episode in a shorter time or fulfil the criteria for a longer time.”*

This reveals an issue at the “application” level of Bloom’s Taxonomy. GPT-4 seemed to remember the fact about the temporal criterion (“remember”) and also understand its concept (“understand”). However, it struggled with the application of this rule, suggesting flexibility where there is conventionally rigidity. This can be interpreted as a misapplication of the learned rule and thus a mistake made at the “application” level of Bloom’s learning hierarchy.

##### 3.2.2.4 Bloom’s Taxonomy: Evaluate

The process of evaluating is found at the fifth level of Bloom’s Taxonomy. It is defined as “making judgments based on criteria and standards”.^8^ GPT-4 made a mistake at Bloom’s “evaluate” level in a more complex question testing the judgement regarding the ideal therapy for a patient. Due to the complexity, the question is presented in full length:

*A 57-year-old ready-to-wear clothing maker is referred to the psychosomatic outpatient clinic by her family doctor because of persistent back pain. She reports “terrible” pain in her lower back. She has been unable to work for seven months because of this and can hardly sleep. No one wanted to help her; the family doctor only sent her from doctor to doctor. (The findings from orthopaedics, neurology, neurosurgery, and radiology that she brought with her show minor disc changes on several levels, but no neurological deficits). She does not want to undergo surgery because “they probably could not earn enough money from her as a patient with public insurance”. Now the family doctor has prescribed her an antidepressant, which she will not take. She still has hope of improvement, but if it continues like this, she does not know whether she will put an end to her life. What is the first course of action indicated?*

*A: Immediate admission to psychiatric ward, against the patient’s will if necessary.*

*B: Initiate inpatient psychosomatic treatment.*

*C: Observation of the symptoms under medication with NSAIDs.*

*D: Obtain a neurosurgical second opinion after current imaging.*

*E: Clarification of secondary disease gain and immediate termination of incapacity for work.*

When we used the short prompt version, GPT-4 chose the answer: Plan a psychosomatic inpatient treatment. In its brief explanation, GPT-4 correctly judged why this was the most likely answer.

However, when we used the detailed prompt, GPT-4 chose the distractor E and sent the patient immediately back to work. GPT-4 argues:

*After careful consideration, answer E is most appropriate, but with modification. A detailed explanation of psychosomatic correlations and the possible role of secondary illness gain might help the patient to better understand her situation and is thus indicated. However, immediate cessation of incapacity should not be forced, as this could potentially exacerbate the patient’s pain and frustration. It would make sense to first seek outpatient psychosomatic treatment to help the patient cope with her pain and focus on long-term recovery.*

In this example, it is evident that GPT-4 knows the facts (“remember”), understands them (“understand”), applies them to this situation (“apply”), and correctly analysis the situation (“analyse”). GPT-4 can even be observed to evaluate but fails at evaluating the consequences of its decision. While GPT-4 seems to be aware of this and thus wants to modify the answer, it incorrectly evaluates the answer options and does not recommend inpatient psychosomatic treatment.

## 4 Discussion

Tools based on LLMs are rapidly evolving and influencing medicine^1^ and medical education.^2^ In this article, we show that GPT-4 yields a high accuracy in answering psychosomatic medicine questions with success rates well above 90%. This confirms results from other groups, which also show high exam scores, e.g., in USMLE^10^ and other board exams.^23^ Although we detected high correct rates for our detailed and short prompt versions, GPT-4 answered more questions correctly when we used the detailed prompt (93% vs. 91%). While our study was not designed to assess reasons for the difference, there are two likely explanations for this difference that deserve attention. First, the difference rates for the two prompt versions could represent a true difference in the LLM’s performance with respect to the prompt version.^24^ Second, the difference could be a random variation that is known to occur even when the same prompt is used more than once.^25,26^

We were further interested in GPT-4’s performance compared to that of medical students. Here, our analysis revealed that questions answered correctly by GPT-4 were significantly easier than questions that were incorrectly answered. This difference could be observed for the detailed and short prompt versions. For further comparison, it has to be noted that the question difficulty is not a fixed/static variable but is dependent and calculated on the basis of the responses from human students.^16^

However, in order to understand why GPT-4 fails at some questions, we further analysed those using a qualitative approach. It is well known that incorrect/inaccurate information is an important issue with LLMs.^27^ To describe the cognitive process underlying learning, Bloom’s Taxonomy has emerged as a frequently used standard.^8^ To the best of our knowledge, the levels at which cognitive errors occur for GPT-4 have not yet been elucidated. We thus performed a detailed assessment of the answers and reasoning GPT-4 provided.

In our analysis, we found that most errors were made at the lowest level of Bloom’s Taxonomy, “remember”. In these answers, GPT-4 failed at naming or using a specific fact, as evident from the text response. In the example presented above, GPT-4 named most of the diagnostic criteria for a somatization disorder but did not mention the time domain. In this context, it is important to note that GPT-4 has been trained with publicly available and licensed data (not specified in detail by OpenAI).^28^ The information required in this example is publicly available in the ICD-10^19^ and thus expected to be included in the GPT-4 training data.

In a recent study, Johnson and colleagues evaluated ChatGPT for its accuracy in medical information.^29^ Using a quantitative approach, they found that GPT-3.5 provided medical answers that were between mostly and almost correct. It is important to note that these results were generated using GPT-3.5, the older model, compared to the model we used, GPT-4 (which is claimed to be “40% more likely to produce factual responses than GPT-3.5”^28^ and shows a better performance in medical exams^23^). However, the results reported by Johnson et al. are in line with our findings: while GPT-4 uses most facts correctly and completely, it sometimes fails at a specific detail. In psychosomatic medicine, we observed this to be a diagnostic criterion. While this can be an issue, too, missing some specific facts in other areas can make all the difference for patient outcomes. Thus, it is important for those using GPT-4 in medicine to keep in mind that specific facts can be wrong and/or missing.

Some errors were found for the second level of Bloom’s Taxonomy, “understand”. While GPT-4 generally showed a good reasoning capability^28^ and errors were sparse, we were surprised that some answers yielded obvious logical flaws, as presented in the example above. In this response, GPT-4 confidently presents subsequent sentences that don’t correlate logically. Mechanistically, language models such as GPT utilize likelihood maximization, generating text based on what most likely follows.^12,30^ However, this approach can result in what is called hallucination: “content that is nonsensical or unfaithful to the provided source content”.^30^ The resulting medical information might sound very confident but be incorrect,^31^ thereby posing a significant threat for medical applications.^5^ This does raise ethical concerns on the use of AI systems for patient-related work, particularly as GPT-4’s algorithms and ethical models are unknown and variable.^4^ Because GPT-4 is not considered sentient, it neither knows nor cares about the accuracy of its responses.

We also detected some mistakes that represented Bloom’s level of “application”. In our representative example, GPT-4 was quite flexible in applying a very strict time criterion (see example above). This can be interpreted in the context of the process of training LLMs. Although little has been published about this process, classification systems probably represent only a small amount of the data available on a certain subject. It can be further assumed that information designed for the public might not be as specific as strict diagnostic criteria because it serves another audience. Following GPT-4’s likelihood maximization approach, LLMs might thus neglect the specific but probably underrepresented piece of information. Furthermore, GPT-4 has been observed to perform poorly at pure calculation tasks,^31^ probably also challenging strictly numerical criteria. This is not entirely surprising, as GPT-4 is a *large language model*, with an emphasis on *language*, and is not intended to be used as a calculator. We found only one mistake for Bloom’s “evaluate” level, in which GPT-4 incorrectly judged a medically complex situation.

Our study has some limitations that deserve discussion. First, we focussed on Bloom’s Taxonomy. While Bloom provided good operationalization for cognitive processes, the taxonomy represents a continuum where more than one level can be activated in a single question.^8^ However, we observed that most errors could be attributed to one cognitive level. Moreover, we chose questions from psychosomatic medicine because many aspects can be covered by written language and do not require images or many numbers. However, psychosomatic medicine is a specialty in which treatment can be individual and especially complex, making categorical judgement harder. Furthermore, our data was acquired once and at a specific date. As the performance of GPT-4 varies over time, this could reduce generalizability.^32^

In summary, we found that GPT-4 performs extremely well on psychosomatic medicine questions. Questions answered correctly by GPT-4 were also easier for human students compared to incorrectly answered questions. When analysing the mistakes GPT-4 made, we found that most errors corresponded to lower-order cognitive levels, such as “remember” and “understand”. While we still found some mistakes for “apply”, very few errors were found for “analyse” and “evaluate” (“create” was not assessed in our study). To the best of our knowledge, we are the first to describe at which cognitive levels GPT-4 makes mistakes in the context of (psychosomatic) medicine.

Our study has important implications. First, GPT-4 is already capable of answering many questions in (psychosomatic) medicine and could thus (given the technology is made available) reduce the effectiveness of summative assessment. Second, GPT-4 sometimes fails with exact facts, correct understanding, and application of knowledge. However, without exact knowledge, these failures are hard to recognize. Thus, output generated by GPT-4 has to be checked for correctness, especially in those domains. Our research can also help in model training, and further studies can use our results to correlate model training and LLM outcomes.

## Data Availability

All data produced in the present study are available upon reasonable request to the authors

